# Brain Aging in Specific Phobia: An ENIGMA-Anxiety Mega-Analysis

**DOI:** 10.1101/2025.03.19.25323474

**Authors:** Kimberly V. Blake, Kevin Hilbert, Jonathan C. Ipser, Laura K.M. Han, Janna Marie Bas-Hoogendam, Fredrik Åhs, Jochen Bauer, Katja Beesdo-Baum, Johannes Björkstrand, Laura Blanco-Hinojo, Joscha Böhnlein, Robin Bülow, Marta Cano, Narcis Cardoner, Xavier Caseras, Udo Dannlowski, Mats Fredrikson, Liesbet Goossens, Hans J. Grabe, Dominik Grotegerd, Tim Hahn, Alfons Hamm, Ingmar Heinig, Martin J. Herrmann, David Hofmann, Hamidreza Jamalabadi, Andreas Jansen, Merel Kindt, Tilo Kircher, Anna L. Klahn, Katja Koelkebeck, Axel Krug, Elisabeth J. Leehr, Martin Lotze, Juergen Margraf, Markus Muehlhan, Igor Nenadić, Wenceslao Peñate, Andre Pittig, Jens Plag, Jesús Pujol, Jan Richter, Isabelle C. Ridderbusch, Francisco Rivero, Axel Schäfer, Judith Schäfer, Anne Schienle, Elisabeth Schrammen, Koen Schruers, Esther Seidl, Rudolf M. Stark, Benjamin Straube, Thomas Straube, Andreas Ströhle, Lea Teutenberg, Sophia I. Thomopoulos, Carlos Ventura-Bort, Renee M. Visser, Henry Völzke, Albert Wabnegger, Julia Wendt, Hans-Ulrich Wittchen, Katharina Wittfeld, Yunbo Yang, Anna Zilverstand, Peter Zwanzger, Lianne Schmaal, Moji Aghajani, Daniel S. Pine, Paul M. Thompson, Nic J.A. van der Wee, Dan J. Stein, Ulrike Lueken, Nynke A. Groenewold

## Abstract

**Introduction:** Specific phobia (SPH) is a prevalent anxiety disorder and may involve advanced biological aging. However, brain age research in psychiatry has primarily examined mood and psychotic disorders. This mega-analysis investigated brain aging in SPH participants within the ENIGMA-Anxiety Working Group.

**Methods:** 3D brain **s**tructural MRI scans from 17 international samples (600 SPH individuals, of whom 504 formally diagnosed and 96 questionnaire-based cases; 1,134 controls; age range: 22-75 years) were processed with FreeSurfer. Brain age was estimated from 77 subcortical and cortical regions with a publicly available ENIGMA brain age model. The brain-predicted age difference (brain-PAD) was calculated as brain age minus chronological age. Linear mixed-effect models examined group differences in brain-PAD and moderation by age.

**Results:** No significant group difference in brain-PAD manifested (**β**_diagnosis_ (SE)=0.37 years (0.43), *p*=0.39). A negative diagnosis-by-age interaction was identified, which was most pronounced in formally diagnosed SPH (**β**_diagnosis-by-age_=-0.08 (0.03), *pFDR*=0.02). This interaction remained significant when excluding participants with anxiety comorbidities, depressive comorbidities, and medication use. Post-hoc analyses revealed a group difference for formal SPH diagnosis in younger participants (22-35 years; **β**_diagnosis_=1.20 (0.60), *p*<0.05, mixed-effects *d* (95% confidence interval)=0.14 (0.00-0.28)), but not older participants (36-75 years; **β**_diagnosis_=0.07 (0.65), *p*=0.91).

**Conclusions:** Brain aging did not relate to SPH in the full sample. However, a diagnosis-by-age interaction was observed across analyses, and was strongest in formally diagnosed SPH. Post-hoc analyses showed a subtle advanced brain aging in young adults with formally diagnosed SPH. Taken together, these findings indicate the importance of clinical severity, impairment and persistence, and may suggest a slightly earlier end to maturational processes or subtle decline of brain structure in SPH.

## Introduction

Specific phobia (SPH) involves fear or anxiety about a specific object or situation that is out of proportion to the actual threat posed (1). SPH is the most prevalent anxiety disorder, usually developing during childhood, with a lifetime prevalence of 2.6% – 12.5% across countries (2,3). Moreover, subclinical fears which have the potential to develop into clinical SPH are a common phenomenon (4). SPH may predict risk for other anxiety or depressive disorders, particularly during the first two decades of life (5–8). Neurobiological research is needed to unravel the biological correlates of aging in SPH, as anxiety disorders are associated with increased risk of early mortality (9). Further, anxiety disorders relate to physiological signs of aging, such as increased oxidative stress, and potential neural markers of aging, such as decreased white matter (WM) and grey matter (GM) density (10). The current study addresses this need by examining relations between SPH and brain aging.

Brain age models inform our understanding of brain health and aging. Such models are generally trained on large datasets of healthy controls (HCs), to learn neurostructural correlates of chronological age (11,12). These models are subsequently applied to unseen datasets to estimate brain age at the individual level. Each participant’s chronological age is subtracted from their brain age, producing the brain predicted age difference (brain-PAD). When an individual’s brain is estimated to be older than their chronological age (reflected in a positive brain-PAD), this may indicate underlying health issues, such as mental illness (13,14). Further, brain age has been associated with an increased risk of all-cause mortality (13). Thus, brain age models may identify unhealthy aging patterns, demonstrating potential to identify risk for psychopathology during aging (13,15).

Most brain age research in psychiatry has so far examined psychotic and mood disorders, with studies finding advanced brain aging in these groups compared to HCs (16–19), and a positive association between brain-PAD and symptom severity (20–23). Further, studies found a potential neuroprotective effect of psychotropic medications (20,24). To the best of our knowledge, only one study has considered brain-PAD in a sample of adults (18-57 years) with anxiety disorders (generalized anxiety disorder, panic disorder, or social anxiety disorder; n=67) compared to HCs (n=65). These patients exhibited a brain-PAD of +2.91 years after correction for antidepressant use (20). Notably, this study did not examine individuals diagnosed with SPH. Few studies have examined abnormalities in brain structure in SPH participants. The largest study to date was recently performed by the ENIGMA-Anxiety (Enhancing NeuroImaging Genetics through Meta-Analysis) Working Group and demonstrated smaller subcortical volumes in the caudate nucleus, putamen, pallidum and hippocampus, and larger thickness in the rostral middle frontal cortex, in SPH participants (n=1,456) compared to HCs (n=2,993; 25), age 5-90 years. However, these correlates only manifested in a subgroup of SPH participants older than age 21 years. Participant age may moderate relations between anxiety disorders and brain age (25,26).

The ENIGMA-Anxiety Working Group is a worldwide research collaboration which performs mega-analyses on large, international multi-site data (27). The primary aim of the present ENIGMA-Anxiety mega-analysis is to compare brain age in a large multi-site sample of adult SPH participants and HCs. We hypothesise that participants with SPH will have a greater brain-PAD than HCs. Further, we examine associations between brain aging in SPH participants and symptom severity. We hypothesise that greater SPH symptom severity relates to a larger brain-PAD.

## Materials and Methods

### Study sample and measures

This multi-site collaborative study included a subset of data (initially n=693 SPH participants, n=1,824 HCs) from 17 research sites participating in the ENIGMA-Anxiety Working Group (25,27), aged 22-75 years (supplementary Figure S1: age density plot). Demographic and clinical information appear in Tables 1a,b. As in a prior report (25), SPH participants were excluded if they had a lifetime diagnosis of psychosis, schizophrenia or bipolar disorder, but retained if they had other comorbidities. HCs were free of all current and past mental disorders.

This secondary data analysis received ethical clearance from the University of Cape Town Human Research Ethics Committee (reference: 675/2021). Participants provided written informed consent during participation with the original studies, and each research site obtained approval from their local ethics committees and institutional review boards to perform the original studies and to share the data with the ENIGMA-Anxiety Working Group.

### Image acquisition and processing

Research sites processed structural T1-weighted magnetic resonance imaging (MRI) images using FreeSurfer (28) and conducted quality control (QC) based on established ENIGMA protocols (https://enigma.ini.usc.edu/; 29) to generate volumes for eight bilateral subcortical regions, surface area and cortical thickness for 34 bilateral regions and total intracranial volume, resulting in 77 features of brain structure in total. Cortical regions were parcellated according to the Desikan-Killiany cortical atlas (30). At the original research sites and central sites, investigators visually inspected the segmentations for failure and poor-quality segmentation, and generated summary statistics, outlier histograms and boxplots. Next, subject-level data was shared with central sites. We excluded participants if they had QC failure for >10% of FreeSurfer regions across both hemispheres (SPH n=82; HC n=116). If participants had <10% missing data, missing values for regions of interest due to QC failure were imputed with the median value of the specific region, by the ENIGMA-MDD brain age model. Data acquired for less than 10 participants per MRI scanner was excluded (n=11 SPH participants from 3 scanners within Protect-AD). Finally, HCs that were part of the sample used to train the ENIGMA-MDD brain age model were excluded from the present study (n=574).

The final sample consisted of 600 SPH participants (current n=335; lifetime and not current n=169; questionnaire-based n=96) and 1,134 HCs from 17 research sites. SPH participants were considered ‘formally diagnosed’ if they were assessed with standardized clinical interviews, and ‘questionnaire-based’ if recruited using questionnaire cut-off scores, primarily used in university and community samples (31–35).

### Brain age calculations

We used a ridge regression brain age model developed by the ENIGMA-MDD Working Group (https://photon-ai.com/enigma_brainage). The model was developed separately in male (n=12,353) and female (n=14,182) HCs aged 18-75 years, using a mega-analytic approach which uses random effects modelling on multi-site, centralized data (18). Specifically, normative models of the association between chronological age and 77 structural brain features were trained with the Python-based *sklearn* package (36) by combining left and right hemisphere FreeSurfer measures and calculating mean volumes of lateral ventricles and subcortical volumes and cortical thickness and surface area (18). This model was validated in a test set of controls by calculating the absolute difference between brain age and chronological age in males (mean absolute error, MAE; standard deviation, SD) = 6.50 (4.91) and females = 6.84 (5.32). The Pearson correlation between chronological age and brain age was *r*=0.85 (*p*<0.001) in the male and *r*=0.83 (*p*<0.001) in the female model. In the current study, brain age was estimated for each participant by using their data for 77 brain structural features as input for the ENIGMA-MDD model (for males or females, respectively).

### Statistical analysis

Statistics were conducted in SPSS version 29 and RStudio 4.2.2. Brain-PAD was calculated by subtracting chronological age from estimated brain age for each participant. The fit of the ENIGMA-MDD brain age model with the data was evaluated by calculating the MAE, Pearson correlation coefficients, and explained variance (R^2^) between chronological age and brain age estimates in the whole sample, and separately in groups (SPH/HC; males/females). Further, model fit was assessed per research site.

The primary research question was tested using a linear mixed-effects model (LME), as implemented in the *nlme* package, 4.2.2 in RStudio. Brain-PAD was the outcome variable, diagnosis (SPH/HC) was the predictor variable, scan site was included as random intercept (in all LMEs), and the mean-centered chronological age, square of the mean-centered chronological age (mean-centered age^2^) and sex were included as covariates (in all LMEs). Diagnosis-by-sex and diagnosis-by-chronological-age interaction terms were added to the model to evaluate whether the brain-PAD is more apparent in specific age groups or in males or females. If these interactions did not significantly influence brain-PAD, they were removed from the model to ensure optimal fit, based on the Akaike and Bayes information criterion (37,38).

Before fitting between-group LMEs, the dataset was checked for normality for brain-PAD, age and cortical thickness (supplementary Figure S2a-S5b). One SPH participant and one HC were identified as outliers due to extreme positive and negative brain-PADs (supplementary Table S1 and Figure S6). Outliers were retained, and results were not impacted significantly by their exclusion (supplementary Table S2).

Certain research sites (FOR2107-MS, Muenster Spider, SHIP, and Teneriffa, Table 1a), differed significantly between groups in mean age and proportion of female participants. Propensity score matching was conducted to match groups (SPH/HC) within these sites on age and sex using the *MatchIt* R package, version 4.5.0 (39). Nearest neighbor matching was implemented as it runs through all case participants and selects the closest eligible control participant for pairing (40). The difference between the propensity scores of each case and control unit was used as the distance measure, and two HCs to one SPH participant ratio was chosen to optimize sample size. The LME was re-run in this matched dataset.

**Table 1a.** Sociodemographic information for each study site included in the main analyses.

| Site | Dx interview/questionnaire cut-off | All |  |  | HC |  |  | SPH |  |  |
| --- | --- | --- | --- | --- | --- | --- | --- | --- | --- | --- |
|  |  | N | % Female | Mean (SD) age | N | % Female | Mean (SD) age | N | % Female | Mean (SD) age |
| **Barcelona** | MINI | 29 | 79.31 | 23.90 (2.06) | 7 | 71.43 | 24.43 (2.44) | 22 | 81.82 | 23.73 (1.96) |
| **Dresden CRC940C5** | CIDI | 96 | 90.63 | 26.68 (5.31) | 44 | 88.64 | 25.86 (5.07) | 52 | 90.31 | 27.37 (5.47) |
| **Dresden SPH subtypes** | CIDI (n=23 cut-off, SNAQ, DFS) | 57 | 75.44 | 25.74 (4.72) | 18 | 66.67 | 25.11 (4.20) | 39 | 79.49 | 26.03 (4.97) |
| **FOR2107 MR** | SCID | 333 | 58.56 | 37.22 (12.55) | 322 | 58.70 | 37.16 (12.55) | 11 | 54.55 | 38.91 (12.98) |
| **FOR2107 MS** | SCID | 170 | 66.47 | 32.58 (12.03) | 146 | 65.75 | 31.66 (11.69)** | 24 | 70.83 | 38.21 (12.86)** |
| **Graz I** | SCID | 55 | 58.18 | 31.76 (10.18) | 26 | 57.69 | 30.04 (8.67) | 29 | 58.62 | 33.31 (11.30) |
| **Graz II** | Other | 31 | 61.29 | 31.84 (11.24) | 15 | 66.67 | 30.27 (10.05) | 16 | 56.25 | 33.31 (12.20) |
| **Greifswald Spider Snake** | Cut-off (SNAQ, SPQ) | 20 | 100.00 | 25.00 (2.13) | 10 | 100.00 | 25.80 (2.15) | 10 | 100.00 | 24.20 (1.87) |
| **Muenster Dental Phobia** | SCID/DAS | 26 | 73.08 | 30.19 (10.80) | 14 | 64.29 | 28.71 (10.33) | 12 | 83.33 | 31.92 (11.52) |
| **Muenster SFBTRR-58 C09** | SCID | 58 | 87.93 | 31.69 (9.63) | NA | NA | NA | 58 | 87.93 | 31.69 (9.63) |
| **Muenster Spider** | SCID | 234 | 56.84 | 38.61 (11.16) | 212 | 53.77* | 39.85 (10.87)** | 22 | 86.36* | 26.64 (5.36)** |
| **Protect-AD** | DSM-5 SPH scale | 34 | 58.82 | 37.15 (11.84) | NA | NA | NA | 34 | 58.82 | 37.15 (11.84) |
| **RepSpi** | Cut-off (SPQ) | 19 | 84.21 | 24.16 (4.95) | 11 | 72.73 | 22.73 (1.10) | 8 | 100.00 | 26.13 (7.32) |
| **SHIP** | CIDI (n=3 cut-off) | 438 | 60.05 | 53.95 (11.64) | 303 | 51.82** | 55.25 (12.02)** | 135 | 78.52** | 51.01 (10.17)** |
| **Teneriffa** | Other | 32 | 71.88 | 35.72 (11.28) | 6 | 50.00 | 25.83 (6.70)* | 26 | 76.92 | 38.00 (10.91)* |
| **Uppsala** | Cut-off (Spider phobia questionnaire) | 40 | 77.50 | 27.05 (7.66) | NA | NA | NA | 40 | 77.50 | 27.05 (7.66) |
| **Wuerzburg SFBTRR-58 C09** | SCID | 62 | 85.48 | 30.45 (7.69) | NA | NA | NA | 62 | 85.48 | 30.45 (7.69) |
| **Total across samples** | NA | 1734 | 65.80 | 38.64 (14.55) | 1134 | 58.82 | 40.42 (15.01) | 600 | 79.00 | 35.28 (13.01) |

Note. Table includes information for the final dataset after exclusions (e.g. due to FreeSurfer quality control failure). CIDI, Composite International Diagnostic Interview; Cut-off, refers to specific phobia participants included based on questionnaire cut-off scores; DAS, Dental Anxiety Scale; DFS, Dental Fear Survey; Dresden CRC940C5: DFG Collaborative Research Centre 940, project C5; Dx, diagnosis; FOR2107 MR: DFG-Research Group 2107 Marburg site; FOR2107 MS: DFG-Research Group 2107 Muenster site; Muenster SFBTRR-58 C09: DFG Collaborative Research Centre Transregio 58, project C09, Muenster site; HC, healthy controls; MINI, Mini International Neuropsychiatric Interview; N, number; NA, not applicable; Protect-AD: Providing Tools for Effective Care and Treatment of Anxiety Disorders consortium, specific phobia sample; SCID, Structured Clinical Interview for DSM; SD, standard deviation; SHIP: Study of Health in Pomerania; SNAQ, Snake Questionnaire; SPH, specific phobia; SPQ, Spider Questionnaire; Wuerzburg SFBTRR-58 C09: DFG Collaborative Research Centre Transregio 58, project C09, Wuerzburg site.\* $p < .05$ ; \*\* $p < .001$ (independent samples T-Test and Chi-square tests).

**Table 1b.** SPH clinical information for each study site included in the main analyses.

| Site | SPH | Comorbidity (lifetime/current) |  | Medication use |  | AOO | STAI-T | Symptom severity centiles | BDI II |
| --- | --- | --- | --- | --- | --- | --- | --- | --- | --- |
|  | % Current, lifetime or cut-off | % ANX | % MDD | % any use | % SSRI/SNRI | Mean (SD) | Mean (SD) | Mean (SD) | Mean (SD) |
| Barcelona | 100.00 current | 0.00/0.00 | 0.00/0.00 | NA | NA | NA | 4.27 (1.55) | 6.82 (1.40) | NA |
| Dresden CRC940C5 | 100.00 current | 15.38/7.69 | 11.54 lifetime | 0.00 | 0.00 | 5.73 (3.92) | 3.17 (1.28) | 7.85 (1.32) | 1.25 (0.62) |
| Dresden SPH subtypes | 41.03 current, 58.97 cut-off | 2.56/10.26 | 0.00/0.00 | 7.69 | 2.56 (n=38 NA) | NA | NA | 8.36 (0.54) | 1.67 (1.31) |
| FOR2107 MR | 100.00 current | 18.18 current | 9.09/72.73 | 63.64 | 45.45 | NA | 7.27 (1.19) | NA | 4.55 (1.29) |
| FOR2107 MS | 83.33 current, 16.67 lifetime | 12.50/25.00 | 4.17/58.33 | 45.83 | 12.50 (n=1 NA) | NA | 6.00 (1.53) | NA | 3.09 (1.34) |
| Graz I | 100.00 current | 0.00/0.00 | 0.00/0.00 | 0.00 | 0.00 | 13.42 (8.26) | NA | 8.69 (1.49) | NA |
| Graz II | 100.00 current | 0.00/0.00 | 0.00/0.00 | 0.00 | 0.00 | 8.06 (4.81) | NA | 8.50 (1.10) | NA |
| Greifswald Spider Snake | 100.00 cut-off | 0.00/0.00 | 0.00/0.00 | NA | NA | NA | NA | 6.80 (1.23) | NA |
| Muenster Dental Phobia | 100.00 cut-off | 0.00/0.00 | 0.00/0.00 | 0.00 | NA | NA | 3.08 (1.62) | 7.42 (1.38) | 1.36 (0.51) |
| Muenster SFBTRR-58 C09 | 100.00 current | 0.00/0.00 | 1.72/1.72 | 43.10 | 0.00 | 6.14 (5.18) | 3.15 (1.42) | 7.76 (0.68) | 1.25 (0.62) |
| Muenster Spider | 100.00 current | 0.00/0.00 | 0.00/0.00 | 0.00 | 0.00 | NA | 4.15 (1.66) | 7.47 (1.65) | 1.23 (0.43) |
| Protect-AD | 100.00 lifetime | 73.53 lifetime | 29.41 lifetime | 0.00 | NA | NA | NA | 4.26 (2.06) | 3.21 (1.67) |
| RepSpi | 100.00 lifetime | 0.00/0.00 | 0.00/0.00 | 0.00 | NA | NA | 3.00 (1.77) | 7.50 (1.31) | NA |
| SHIP | 0.74 current, 97.04 lifetime, 2.22 cut-off | 17.04 lifetime | 28.15 lifetime | 22.22 (n=3 NA) | 8.12 | 14.26 (11.34) | NA | NA | 2.02 (1.40) |
| Teneriffa | 100.00 current | 0.00/0.00 | 0.00/0.00 | 0.00 | 0.00 | NA | NA | 7.31 (1.12) | NA |
| Uppsala | 100.00 cut-off | 0.00/0.00 | 0.00/0.00 | 0.00 | NA | 5.74 (2.80) | NA | 6.78 (1.03) | NA |
| Wuerzburg SFBTRR-58 C09 | 100.00 current | 0.00/0.00 | 1.61 lifetime | 40.32 | 0.00 | 7.79 (3.83) | 3.27 (1.54) | 8.00 (0.81) | 1.26 (0.51) |
| Total across all samples | 55.83 current, 28.17 lifetime, 16.00 cut-off | 10.00/2.67 | 9.67/3.83 | 16.83 | 7.67 (n=165 NA) | 9.62 (8.48) | 3.77 (1.83) | 7.45 (1.61) | 1.84 (1.33) |

Note. ANX, anxiety; AOO, age of onset; BDI, Beck Depression Inventory II; Dresden CRC940C5: DFG Collaborative Research Centre 940, project C5; FOR2107 MR: DFG-Research Group 2107 Marburg site; FOR2107 MS: DFG-Research Group 2107 Muenster site; Muenster SFBTRR-58 C09: DFG Collaborative Research Centre Transregio 58, project C09, Muenster site; MDD, major depressive disorder; NA, not applicable; Protect-AD: Providing Tools for Effective Care and Treatment of Anxiety Disorders consortium, specific phobia sample; SD, standard deviation; SHIP: Study of Health in Pomerania; SNRI, serotonin and norepinephrine reuptake inhibitor; SPH, specific phobia; SSRI, selective serotonin reuptake inhibitor; STAI-T, State-Trait Anxiety Inventory – Trait; Wuerzburg SFBTRR-58 C09: DFG Collaborative Research Centre Transregio 58, project C09, Wuerzburg site.

We repeated the main LME in SPH participants with a formal current and lifetime SPH diagnosis only, excluding those classified as SPH solely based on questionnaire cut-off scores (Table 1a). We repeated the analysis in formally diagnosed SPH participants with current SPH, as compared to HCs, excluding two research sites (Protect-AD, RepSpi) which reported lifetime SPH only. Next, an exploratory analysis was run in the research sites with both SPH and HC participants, excluding those without HCs, to account for potential site-effects. Subgroup analyses were conducted comparing the brain-PAD in unmedicated SPH participants and HCs, and as a supplementary analysis, in medicated SPH participants and HCs. Two comorbidity analyses were run, in which SPH participants with comorbid MDD were excluded, and with any comorbid anxiety disorder were excluded (generalized anxiety disorder, social anxiety disorder, panic disorder, agoraphobia, separation anxiety, and unspecified anxiety disorder). False discovery rate (FDR) correction was applied to the n=5 sub-analyses of interest, specifically, formal current and lifetime SPH, current SPH, unmedicated SPH, no anxiety comorbidity, and no MDD comorbidity, with a significance threshold set at *p*<0.05. Finally, a post-hoc analysis was conducted where participants were split by median age of 35 years (22-35 years; 36-75 years), allowing for an approximately even sample size per age bin and maximum statistical power. The main LME was rerun in these age bins and repeated in formally diagnosed SPH and HCs within these age bins.

The second hypothesis, namely that greater symptom severity would predict greater brain-PAD in SPH, was tested using an LME conducted in SPH participants only. Due to different symptom severity measures used across research sites, SPH participants were classified into severity centiles, in line with prior work on this sample (25; present study formally diagnosed SPH mean centile score = 7.49 (1.70); questionnaire-based mean centile score = 6.19 (2.72)). Brain-PAD was the outcome variable, SPH symptom severity was the predictor variable, scanner site was included as a random intercept, and mean-centered age, mean-centered age^2^ and sex were included as covariates. This same model was run again with STAI-Trait (41) as the predictor variable. Analyses were repeated in formally diagnosed SPH participants (lifetime and current) only.

## Results

### Model fit

The ENIGMA-Anxiety SPH dataset fit well with the ENIGMA-MDD brain age model, with comparable MAE (full sample = 7.26 (SD=5.41), SPH = 7.43 (5.81), HC = 7.17 (5.19)), MAE weighted by sample age range (weighted MAE = 0.14 in full sample and per group) and Pearson correlations (full sample = 0.80, SPH = 0.76, HC = 0.81) between brain age and age to those observed in Han et al. (18), using the same model (supplementary Table S3-S5: model fit per study site).

### Brain-PAD in SPH versus HCs

SPH participants (n=600) had a mean chronological age of 35.28 (13.01), brain age of 39.18 (11.28) years, and thus mean brain-PAD of 3.90 (8.59) years. HCs (n=1134) had a mean chronological age of 40.42 (15.01), brain age of 41.33 (12.76) years and mean brain-PAD of 0.91 (8.80) years. This brain-PAD did not differ significantly between SPH participants and HCs, after adjusting for scanner site, mean-centered age, mean-centered age^2^ and sex (Table 2; supplementary Figure S7: brain-PAD residuals). A significant main effect of (mean-centered) chronological age was observed (Table 2). After including an a priori diagnosis-by-age interaction term in the model, we found a main effect of (mean-centered) age, as well as an effect of mean-centered age^2^ and a significant interaction of diagnosis-by-age (Table 2; Figure 1; supplementary Figure S8 shows scatter plots per site).

**Figure 1.**
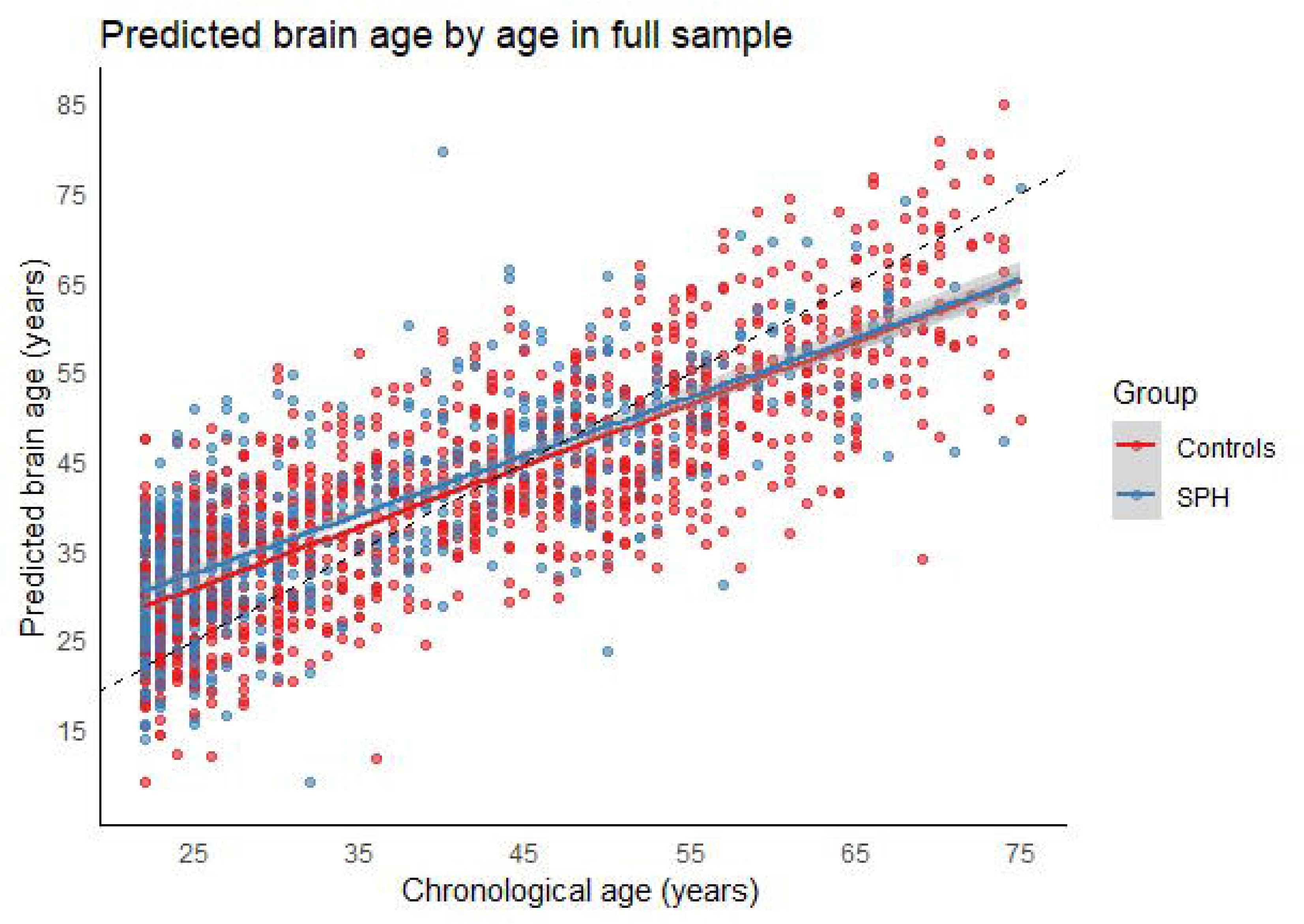
Chronological age plotted against predicted brain age in the full sample. Note. SPH, specific phobia. Diagonal dashed line represents the line of identity (x = y).

**Table 2.** Between-group differences (SPH vs HC) in brain-PAD, diagnosis-by-age interaction.

| GLM without interaction term | $\beta$ (SE) | t-value | p | Cohen's d, (95% CI) |
| --- | --- | --- | --- | --- |
| Intercept | 1.89 (0.98) | 1.93 | 0.05 |  |
| Diagnosis | 0.42 (0.43) | 0.99 | 0.32 | 0.05 (-0.05-0.14) |
| Sex | 0.20 (0.37) | 0.55 | 0.59 |  |
| AgeC | -0.39 (0.02) | 22.02 | < 0.001 |  |
| AgeC<sup>2</sup> | -0.00 (0.00) | -1.66 | 0.10 |  |
| GLM with interaction term | $\beta$ (SE) | t-value | p | |
| Intercept | 1.83 (1.00) | 1.83 | 0.07 |  |
| Diagnosis | 0.37 (0.43) | 0.86 | 0.39 | 0.04 (-0.05-0.13) |
| Sex | 0.22 (0.37) | 0.60 | 0.55 |  |
| AgeC | -0.37 (0.02) | -19.18 | < 0.001 |  |
| AgeC<sup>2</sup> | -0.00 (0.00) | -2.04 | 0.04 |  |
| Diagnosis-by-ageC | -0.07 (0.03) | -2.45 | 0.02 |  |

Note: AgeC, mean-centered chronological age; AgeC<sup>2</sup>, mean-centered chronological age squared; CI, confidence intervals; Diagnosis-ageC, diagnosis-by-age-centered interaction term; GLM, general linear model; SE, standard error. $\beta$ is measured in years.

Subsequently, diagnosis-by-age-squared and diagnosis-by-sex interaction terms were added to new models, one at a time. However, neither interaction term was significant (**β**_diagnosis-by-age_^2^=-0.00 (0.00), *t*-value=-0.80, *p*=0.42; **β**_diagnosis-by-sex_=-0.70 (0.83), t-value=-0.85, *p*=0.40), and both were removed to ensure optimal model fit (lower Akaike and Bayes information criterion: AIC=11733.62, BIC=11771.81).

An exploratory LME was run in research sites including both SPH (n=480) and HC participants (n=1,134), to ensure the significant diagnosis-by-age interaction was not explained by site effects. Despite a smaller sample size, the diagnosis-by-age interaction remained a similar magnitude, and approached statistical significance in this model (**β**_diagnosis-by-age_=-0.06 (0.03), *p*=0.06; supplementary Table S6), confirming that site effects do not explain the interaction. Finally, following nearest neighbor propensity score matching (supplementary Table S7, Figure S9); the brain-PAD did not significantly differ between SPH participants and HCs, while the diagnosis-by-age interaction remained significant (Table S8).

### Clinical subgroup analyses

To assess if the diagnosis-by-age interaction was upheld in subgroups, post-hoc sensitivity analyses were conducted. First, the primary LME was re-run in formally diagnosed SPH participants compared to HCs (Table 3; supplementary Table S9-S10: clinical information). The results were consistent with those from the model using all SPH participants, with no significant difference in brain-PAD between groups. Notably, the significant diagnosis-by-age interaction effect was larger than in the full sample, and survived FDR correction. The interaction remained significant after FDR correction when the LME was conducted in current SPH only (without lifetime; Table 3). A subgroup analysis did not reveal an effect of diagnosis when comparing brain-PAD between unmedicated SPH participants and HCs, however, the diagnosis-by-age interaction remained significant after FDR correction (Table 3). Two additional subgroup analyses were conducted in SPH participants without anxiety comorbidities (versus HCs), and in SPH participants without MDD comorbidity (versus HCs; Table 3). Brain-PAD did not differ significantly between groups, however, the diagnosis-by-age interaction remained significant after FDR correction, and therefore could not be explained by comorbid anxiety, comorbid MDD or medication use.

**Table 3.** Between group differences in brain-PAD, diagnosis and diagnosis-by-age interaction parameters.

|  | SPH | HC | Dx |  |  |  | Dx-by-ageC |  |  |  |
| --- | --- | --- | --- | --- | --- | --- | --- | --- | --- | --- |
| | n | n | $\beta$ (SE) | t-value | p | Cohen's d (95% CI) | $\beta$ (SE) | t-value | p | pFDR |
| SPH (no medication use) vs HCs | 463 | 1,134 | 0.03 (0.47) | 0.07 | 0.94 | 0.00 (-0.09-0.10) | -0.07 (0.03) | -2.11 | 0.03 | 0.03 |
| SPH (no ANX comorbidity) vs HCs | 515 | 1,134 | 0.09 (0.45) | 0.19 | 0.85 | 0.01 (-0.09-0.11) | -0.07 (0.03) | -2.35 | 0.02 | 0.02 |
| SPH (no MDD comorbidity) vs HCs | 495 | 1,134 | 0.38 (0.47) | 0.80 | 0.42 | 0.04 (-0.06-0.14) | -0.08 (0.03) | -2.51 | 0.01 | 0.02 |
| Diagnosed SPH (lifetime, current) vs HCs | 504 | 1,134 | 0.51 (0.44) | 1.15 | 0.25 | 0.06 (-0.04-0.15) | -0.08 (0.03) | -2.85 | 0.00 | 0.02 |
| Diagnosed SPH (current) vs HCs | 335 | 1,134 | 0.26 (0.64) | 0.40 | 0.69 | 0.02 (-0.08-0.13) | -0.11 (0.05) | -2.41 | 0.02 | 0.02 |

Note: ANX, anxiety; CI, confidence intervals; Dx, diagnosis; Dx-by-ageC, diagnosis-by-age-centered interaction term; FDR, false discovery rate; HC, healthy controls; MDD, major depressive disorder; n, number; SE, standard error; SPH, specific phobia. $\beta$ is measured in years.

For completeness, supplementary analyses were conducted in medicated SPH vs HCs (supplementary S12), and in formally diagnosed SPH (lifetime, current) vs HCs, in only sites with both SPH and HC groups (Supplementary Table S11; **β**_diagnosis_=0.49 (0.47), *p*=0.30, Cohen’s *d* (95%CI)=0.06 (−0.05-0.16); **β**_diagnosis-by-age_=-0.07 (0.03), *p*=0.02).

### Post-hoc: median split age

The consistently observed diagnosis-by-age interaction was explored further by running the primary LME in different age groups, split by median age (35 years) (supplementary Table S13-S14: group clinical and demographic information; supplementary Figure S10a-S10b: brain age scatterplots). A difference in brain-PAD of almost 1 year was observed between SPH participants and HCs in the younger age group, however this was not significant (Table 4). No difference in brain-PAD was observed between SPH participants or HCs in the older age group (Table 4). Due to the most pronounced diagnosis-by-age interaction effect found in formally diagnosed SPH participants (current and lifetime) compared to HCs, the post-hoc analysis was repeated in this group. A significantly different brain-PAD of +1.20 years was observed in SPH participants compared to HCs in the younger age group (*p*=0.047), but not the older age group (Table 4).

**Table 4.** Between group differences in brain-PAD, diagnosis parameters.

|  | **SPH n** | **HC n** | **$\beta$ (SE)** | **t-value** | ***p*** | **Cohen's *d* (95% CI)** |
| --- | --- | --- | --- | --- | --- | --- |
| **All SPH vs HCs, 22-35 yrs** | 355 | 532 | 0.94 (0.57) | 1.66 | 0.10 | 0.11 (-0.02-0.24) |
| **All SPH vs HCs, 36-75 yrs** | 245 | 602 | 0.01 (0.64) | 0.02 | 0.99 | 0.00 (-0.13-0.14) |
| **Diagnosed SPH vs HCs, 22-35 yrs** | 272 | 532 | 1.20 (0.60) | 1.99 | **0.05** | 0.14 (0.00-0.28) |
| **Diagnosed SPH vs HCs, 36-75 yrs** | 232 | 602 | 0.07 (0.65) | 0.11 | 0.91 | 0.01 (-0.13-0.14) |

Note: CI, confidence intervals; n, number; HC, healthy controls; SE, standard error; SPH, specific phobia; vs, versus; yrs, years. $\beta$ is measured in years.

### Symptom severity analyses

SPH symptom severity centile score and STAI-T score were not significantly associated with brain-PAD in SPH participants (symptom severity centile score: n=427, **β**_symptom-severity_=0.18 (0.24), t-value=0.74, *p*=0.46; STAI-T score: n=266, **β**_STAI-T_=0.13 (0.25), t-value=0.54, *p*=0.59). When conducting analyses in formally diagnosed SPH participants only, P-values were slightly larger but still non-significant (symptom severity centile score: n=334, **β**_symptom-severity_=0.19 (0.26), t-value=0.72, *p*=0.47; STAI-T score: n=246, **β**_STAI-T_=0.17 (0.26), t-value=0.64, *p*=0.52).

## Discussion

This multi-site mega-analysis examined whether brain-PAD differed between adult SPH participants and HCs. Brain-PAD did not relate to SPH in the full sample, or when limiting the analyses to SPH participants with a formal diagnosis (vs. HCs). However, a significant diagnosis-by-age interaction persisted across sensitivity analyses, and was most evident in formally diagnosed SPH participants vs HCs. For the purposes of interpretation, post-hoc analyses were conducted, splitting participants by median age. This showed a subtle advanced brain-PAD in younger formally diagnosed SPH participants compared to HCs of +1.20 (0.60) years, Cohen’s *d*=0.14 (age: 22-35 years), but not in older participants (age: 36-75 years). No associations were identified between brain-PAD and symptom severity scores in SPH participants. The diagnosis-by-age interaction suggests a slightly earlier end of maturational processes, or an advanced early decline in neurostructural measures.

### Brain aging patterns according to age

The brain-PAD is correlated with chronological age. Brain age models may underestimate age in older samples and overestimate age in younger samples, particularly when participant age deviates from the training sample mean age (17,42). Including age (linear and quadratic) in the LMEs can correct for this age bias as effectively as applying correction to the brain-PAD metric (43,44). Still, brain age prediction accuracy may be impacted by differing age ranges of the training and test samples (18,45). Here, the model training and study samples had a similar age distribution. Further, the model demonstrated reasonable fit with the present dataset (18). After adjusting for age in analyses, a greater effect may still be expected in older individuals, due to accelerated aging during older age (17). However, the present study identified a positive brain-PAD in younger formally diagnosed SPH participants compared to controls, but not in older participants, suggesting accentuated structural brain aging in young adults with SPH.

Biological and lifestyle mechanisms may impact biological aging. Biological factors, such as oxidative stress, contribute to premature cell death and have been associated with both biological aging and anxiety disorders (46,47), but these may be more relevant in older individuals. Maturation of brain regions such as the amygdala, hippocampus and prefrontal cortex in youth may be vulnerable to unhealthy lifestyle factors, including chronic stress (48,49), such as that experienced in SPH (50). Given evidence of ongoing brain maturation throughout the first three decades of life (51–53), the diagnosis-by-age interaction may be indicative of a slightly earlier end of brain maturation or a subtle ‘advanced’ early decline in structural brain measures.

The size of the brain-PAD observed in the younger formally diagnosed SPH group is smaller than observed previously in a multi-site study of adult clinically diagnosed anxiety disorder participants (+2.91 years after correcting for antidepressant use; 20). However, Han et al. (20) did not assess SPH participants. Further, the group difference in brain-PAD is similar in size to MDD (approximately +1 year; 18,54), and smaller than bipolar disorder and psychotic disorders (+2 years and +3 years respectively; 16,17). The small effect identified in the present study is in line with research showing that structural brain differences in anxiety disorders tend to be subtle (26,55). Although, a recent mega-analysis in a larger version of the present sample suggests that SPH structural effect sizes may exceed that reported in other anxiety disorders (25).

### Clinical subgroups and symptom severity

An advanced brain-PAD was subtly present in younger formally diagnosed SPH, but not in the full age range of formally diagnosed SPH. The formally diagnosed SPH subgroup may present with more clinically severe and persistent disorder than questionnaire-based participants, who were primarily recruited from community and university settings. The questionnaires did not assess significant impairment or disorder persistence (e.g. minimum duration of 6 months), whereas clinical interviews include these criteria (1,56,57). Further, formally diagnosed SPH had a slightly higher mean symptom severity centile score than questionnaire-based SPH. Therefore, it may be expected that advanced brain aging is most pronounced in formally diagnosed participants.

Research has suggested a potential subtle neuroprotective effect of antidepressant use against brain aging (20). The present study found no significant difference in brain-PAD between either medicated or unmedicated SPH and controls. However, only a relatively small percentage of included SPH participants used selective serotonin reuptake inhibitors (SSRIs) or serotonin and norepinephrine reuptake inhibitors (Table 1b), possibly due to limited evidence that SSRIs are beneficial in SPH (58). Potential increased disorder severity in participants taking psychotropic medications could offset possible neuroprotective effects. This aligns with our sub-analysis in medicated SPH participants compared to HCs, where a group difference in brain-PAD of +1.10 years was identified (supplementary Table S12), although this did not reach significance.

SPH patients commonly have comorbid psychopathologies, which may become more severe than the phobia (4,59), highlighting the importance of examining whether comorbidities impact the diagnosis-by-age interaction. We found no significant differences between SPH participants without anxiety disorder comorbidities and HCs, or between SPH participants without MDD comorbidity and HCs. The diagnosis-by-age interaction term remained in both models, indicating that the age-specific relationship between diagnosis and brain-PAD cannot be explained by comorbid anxiety and MDD diagnoses.

Previous studies have found a higher brain-PAD with increased symptom severity, primarily in psychotic disorders (21,22,60). Further, one study detected an association between higher brain-PAD and higher anxiety and depressive symptoms across individuals diagnosed with anxiety disorders, MDD and HCs (20). The present study did not detect an association between brain-PAD and SPH symptom severity centile scores or STAI-T scores (measuring trait anxiety; 41) in the SPH group. The lack of significant relationships between clinical symptoms and brain structure aligns with the finding of a recent ENIGMA-Anxiety study, from which the present study sample was derived (25). Heterogeneity in clinical measures and smaller sample sizes (compared to the primary analysis), may have resulted in decreased sensitivity to detect associations.

### Study limitations and strengths

This is the first mega-analysis to investigate brain aging in a large, international sample of SPH participants. Multisite collaborations, such as ENIGMA, have the major benefit of large sample sizes which increase statistical power to detect group differences (27,61). Further, MRI data pre-processing and quality checking was harmonized across research sites, enhancing methodological homogeneity. Finally, the brain age model was trained on a large independent sample and fit well with the present dataset, demonstrating similar fit statistics to Han et al. (18,54).

Some limitations of the present study deserve to be mentioned. Clinical information was not prospectively harmonized across research sites and not available for all sites, resulting in smaller sample sizes in sub-analyses and decreased power to detect group differences and associations. In addition, not all research sites had control participants. However, supplementary analyses demonstrated that the diagnosis-by-age finding was not impacted by site-effects.

Furthermore, while the present brain age model is sensitive to the effects of aging due to the inclusion of cortical thickness measures (62), it predicts age based on the whole brain. Certain models can estimate brain age for specific brain regions, providing additional information on regional brain aging patterns (19). Caution is important when interpreting findings relating to advanced brain aging or maturation, as underlying mechanisms remain to be elucidated (63). Finally, anxiety disorders commonly start early in life (2), therefore, brain age studies in children and adolescents with anxiety disorders could provide clarity regarding brain maturation and the direction of the relationship between brain aging and anxiety disorders.

## Conclusion

This ENIGMA-Anxiety mega-analysis did not identify significantly advanced brain aging in the full sample of SPH participants. However, a diagnosis-by-age interaction was present across analyses, and was particularly evident in formally diagnosed SPH participants compared to HCs. Further, a subtle advanced brain aging was identified in post-hoc analyses in formally diagnosed young adults with SPH. This finding potentially indicates a slightly earlier end to maturational processes or an advanced decline in structural brain measures. The findings in the present study suggest that advanced brain aging may be more apparent in formally diagnosed participants, indicating the importance of clinical severity, impairment and persistence. Research is needed in younger participants and on regional brain age patterns in SPH to provide clarity on which developmental or aging processes may be implicated by brain age models.

## Data Availability

The ENIGMA-Anxiety Working Group is open to sharing the data and code from this investigation to researchers for secondary data analysis. To request access to volumetric, clinical, and demographic data, an analysis plan can be submitted to the ENIGMA-Anxiety Working Group (http://enigma.ini.usc.edu/ongoing/enigma-anxiety/). Data access is contingent on approval by PIs from contributing samples.

The results of this project were presented in the form of a poster presentation at Society of Biological Psychiatry in Austin, Texas, on 09 May 2024, and at the Southern African Neuroscience Society in Durban, South Africa, on 19 July 2024. The current manuscript is substantively different from the previous work, as it is a full archival report, rather than a poster presentation or an abstract

Blake, K. V., Hilbert, K., Ipser, J. C., Han, L. K., Bas-Hoogendam, J. M., Aghajani, M.,… & Groenewold, N. A. (2024). 67. An Enigma-Anxiety Mega-Analysis Investigating Brain Ageing in Specific Phobia. *Biological Psychiatry*, *95*(10), S126-S127.

## Acknowledgements

The ENIGMA Anxiety Working Group acknowledges the NIH Big Data to Knowledge (BD2K) award for foundational support and consortium development (U54 EB020403 to PMT). For a complete list of ENIGMA-related grant support please see here: http://enigma.ini.usc.edu/about-2/funding/. KVB was supported by the Oppenheimer Memorial Trust (OMT Ref. 22148/01) and Boehringer Ingelheim Fonds. JMBH received funding from NWO (Rubicon 019.201SG.022), Medical Delta (Talent Acceleration Call 2021) and the Dutch Research Agenda (NWA/NeurolabNL NWA.1418.22.025). UD was funded by the Interdisciplinary Center for Clinical Research (IZKF) of the medical faculty of Münster (grant Dan3/022/22 to UD). MF was supported by the Swedish Research Council and the Swedish Brain Foundation. AS was funded by the Bundesministerium für Bildung und Forschung. KS was funded by the Weijerhorst Foundation. DSP was supported by NIMH Intramural Research Award ZIA-MH002781. NAG was supported in part by a grant from Carnegie Corporation of New York. The statements made and views expressed are solely the responsibility of the author. This work was further supported by multiple grants from the German Research Foundation (DFG): to the CRC940/2 (project C5 to KBB and MM), to the FOR2107 (KI588/14-1, and KI588/14-2, and KI588/20-1, KI588/22-1 to TK; NE2254/1-2, NE2254/2-1, NE2254/3-1, NE2254/4-1 to IN; DA1151/5-1, DA1151/5-2, DA1151/9-1, DA1151/10-1, DA1151/11-1 to UD), to the SFB-TRR58 (projects C09 and Z02 to UD and UL). The German research consortia PROTECT-AD (“Providing Tools for Effective Care and Treatment of Anxiety Disorders”: PI: HUW) was funded by the German Federal Ministry of Education and Research (BMBF; www.fzpe.de; grant 01EE1402). MRI data were collected as part of project P4 (PIs: TK and BS; project no. 01EE1402E). Clinical recruitment and data collection was conducted in project P1 (01EE1402A). SHIP is part of the Community Medicine Research net of the University of Greifswald, Germany, which is funded by the Federal Ministry of Education and Research (grants no. 01ZZ9603, 01ZZ0103, and 01ZZ0403), the Ministry of Cultural Affairs and the Social Ministry of the Federal State of Mecklenburg-West Pomerania. MRI scans in SHIP and SHIP-TREND have been supported by a joint grant from Siemens Healthcare, Erlangen, Germany and the Federal State of Mecklenburg-West Pomerania. This study was further supported by the German Research Foundation (GR 1912/5-1).

The authors would like to thank Sophia Seemann, Laurenz Endl, Kassandra Friebe, Angela Doss, Petra Bartel and Markus Otto for their help in organizing the data and supporting the quality control process.

