## Supplementary Information for "Brain Aging in Specific Phobia: An ENIGMA-Anxiety Mega-Analysis"

**Supplementary Materials**

**List of Tables and Figures**

Figure S1. Age density plot per diagnostic group.

Figure S2a-S2b. Age boxplots.

Figure S3a-S3b. Brain-PAD boxplots.

Figure S4a-S4b. MAE boxplots.

Figure S5a-S5b. Cortical thickness boxplots.

Table S1. Brain-PAD extreme values.

Figure S6. Boxplot of brain-PAD extreme values.

Table S2. Between-group differences (SPH n=599 vs HC n=1,133) in brain-PAD (outlier removal).

Table S3. Model fit statistics, full sample, and per diagnostic group and sex.

Table S4. Mean absolute error per study site and diagnostic group

Table S5. Pearson’s R and R^2^: brain age and age and brain-PAD and age, per study site and diagnostic group.

Figure S7. Brain-PAD residuals plot for primary linear mixed-effects model without interaction term.

Figure S8. Brain age by age scatterplots per diagnostic group and research site.

Table S6. Between-group differences (SPH n=480 vs HC n=1,134) in brain-PAD, sites with HCs.

Table S7. Summary of balance for matched and unmatched data per study site.

Figure S9. Density plots for age and sex (0 = male; 1 = female), per matched research site.

Table S8. Between-group differences (SPH n=577 vs HC n=766) in brain-PAD, matched dataset.

Table S9. Clinical information for formally diagnosed SPH participants (n=504).

Table S10. Clinical information for questionnaire cut-off SPH participants.

Table S11. Between-group differences in brain-PAD (formally diagnosed SPH, lifetime and current n=384 vs HCs n=1,134) in sites with controls vs HCs.

Table S12. Between-group differences in brain-PAD (medicated SPH n=101 vs HC n=1,134).

Table S13. Demographic and clinical information, participants ≤ 35.

Table S14. Demographic and clinical information, participants > 35.

Figure S10a. Scatterplot of brain age by age in younger formally diagnosed SPH and HCs.

Figure S10b. Scatterplot of brain age by age in older formally diagnosed SPH and HCs.

**Age density plot.**
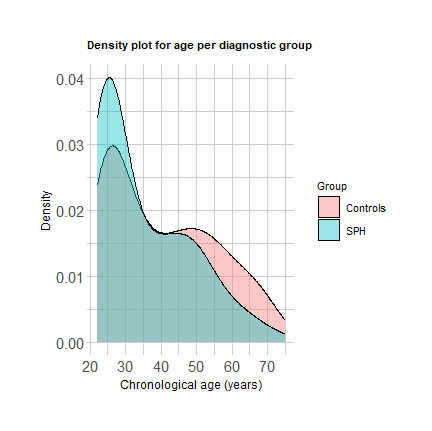

Figure S1. Age density plot per diagnostic group (SPH n=600, HC n=1,134). Note. HC, healthy controls; SPH, specific phobia.

**Tests for normality.**

**Age boxplots.**
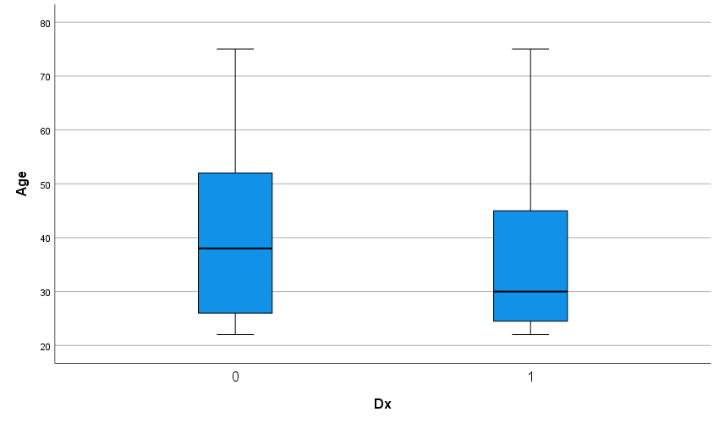

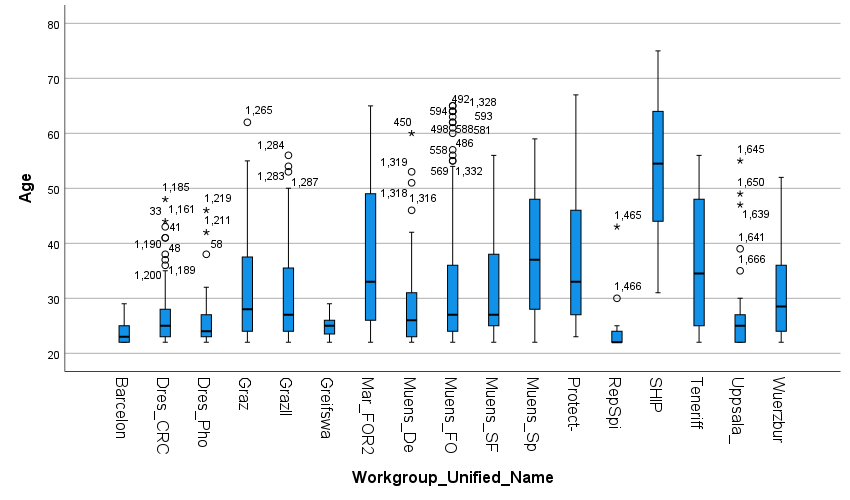

Figure S2a. Age boxplots per diagnostic group. Figure S2b. Age boxplots per research site. Note. Dx, diagnosis. Healthy controls = 0; Specific phobia = 1.

**Brain-PAD boxplots.**
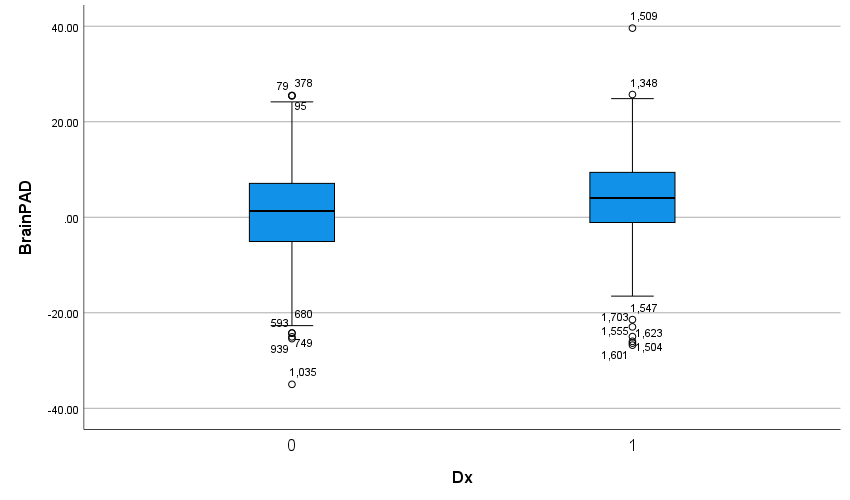

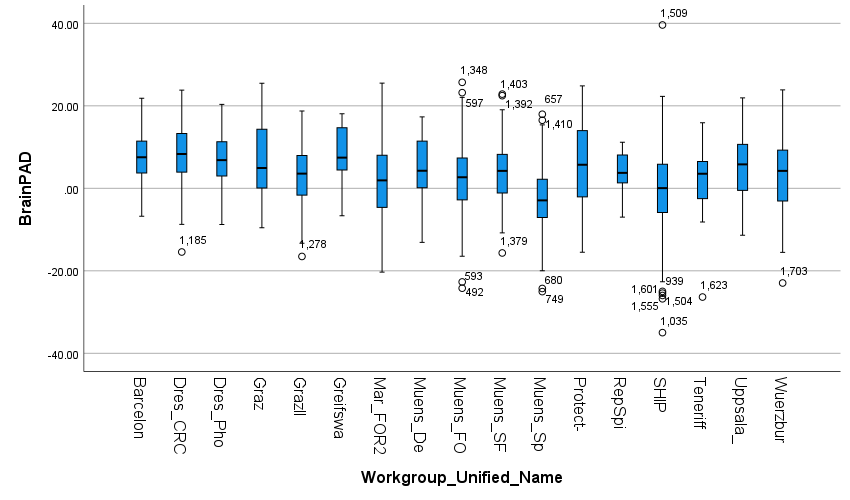

Figure S3a. Brain-PAD boxplots per diagnostic group. Figure S3b. Brain-PAD boxplots per research site. Note. BrainPAD, brain predicted age difference; Dx, diagnosis. Healthy controls = 0; Specific phobia = 1.

**MAE boxplots.**
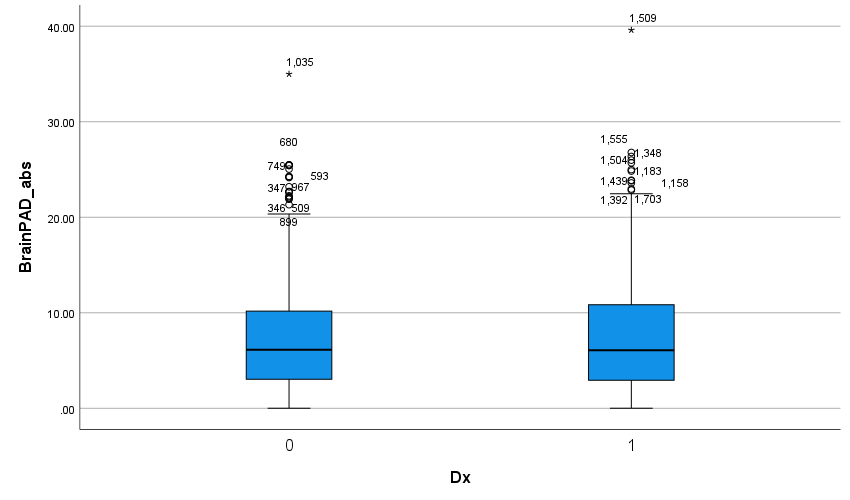

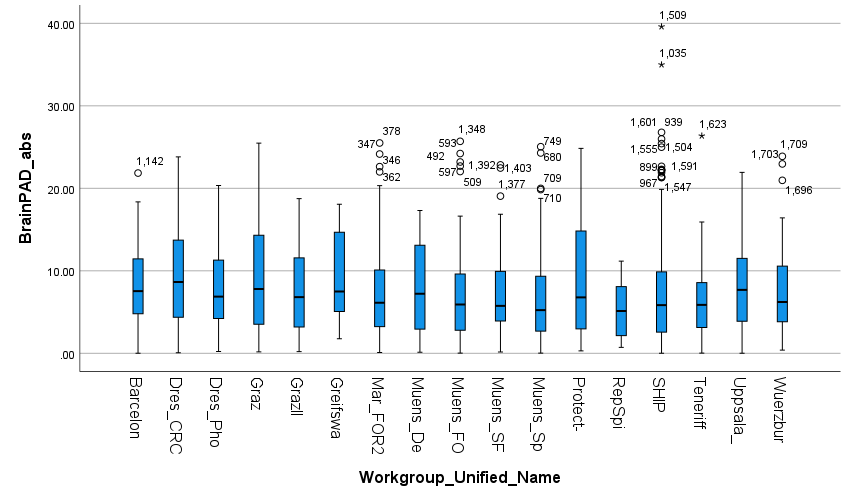

Figure S4a. MAE boxplots per diagnostic group. Figure S4b. MAE boxplots per research site. Note. BrainPAD abs, brain predicted age difference absolute value/mean absolute error; Dx, diagnosis; MAE, mean absolute error. Healthy controls = 0; Specific phobia = 1.

**Cortical thickness boxplots.**
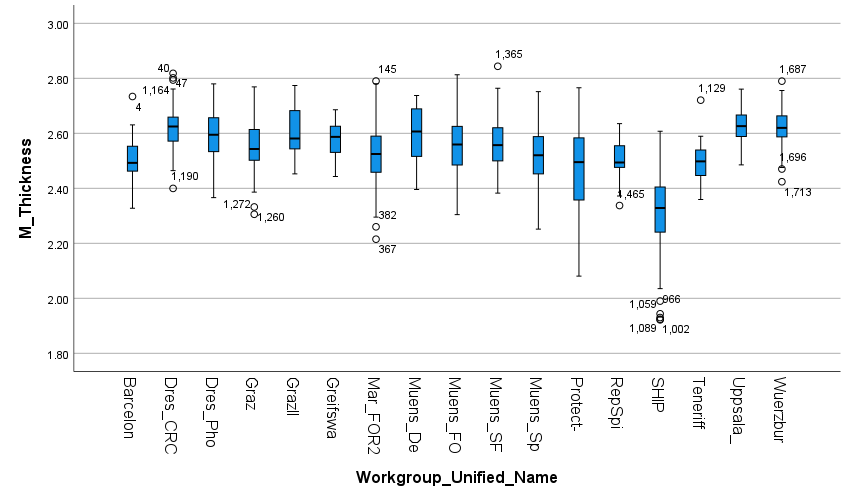

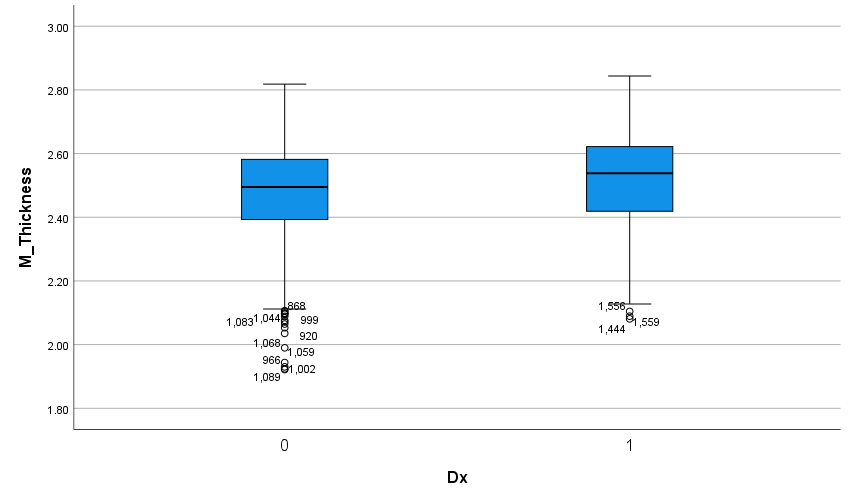

Figure S5a. Cortical thickness boxplots per diagnostic group. Figure S5b. Cortical thickness boxplots per research site. Note. Dx, diagnosis; M Thickness, mean cortical thickness. Healthy controls = 0; Specific phobia = 1.

**Brain-PAD extreme values.**

The below table and boxplot demonstrate the highest and lowest extreme brain-PAD values, generated with SPSS.

Table S1. Brain-PAD extreme values.

|  | Case number | Site | Brain-PAD value |
| --- | --- | --- | --- |
| Highest | 1503 | SHIP | 39.60 |
| Lowest | 1372 | SHIP | -34.99 |

Note. Brain-PAD, brain predicted age difference.

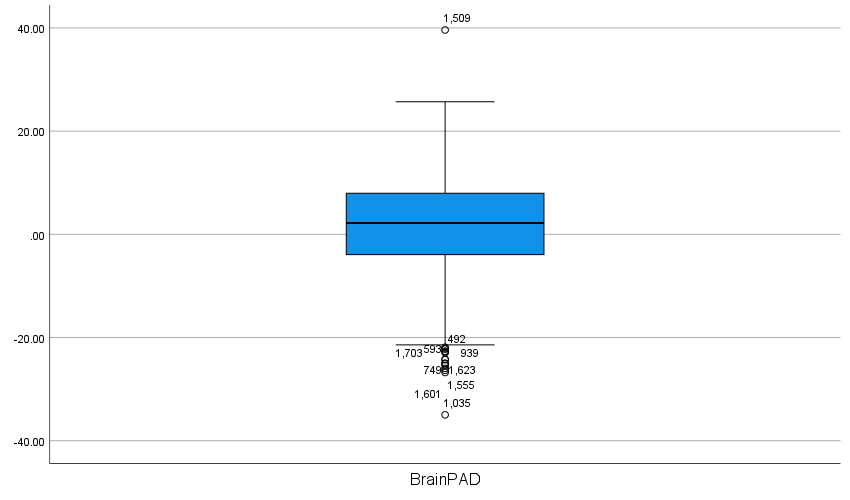

Figure S6. Boxplot of brain-PAD extreme values. Note. BrainPAD, brain predicted age difference.

**Linear mixed-effects model: Brain-PAD between groups with outlier removal (1 SPH; 1 HC).**

Table S2. Between-group differences (SPH n=599 vs HC n=1,133) in brain-PAD (outlier removal)

| LME without interaction term | 𝛃 (SE) | t-value | *p* |
| --- | --- | --- | --- |
| Intercept | 1.90 (0.98) | 1.94 | **0.05** |
| Diagnosis | 0.33 (0.42) | 0.78 | 0.44 |
| Sex | 0.20 (0.36) | 0.55 | 0.58 |
| AgeC | -0.39 (0.02) | -22.16 | ***p*<0.001** |
| AgeC^2^ | -0.00 (0.00) | -1.34 | 0.18 |
| LME with interaction term | 𝛃 (SE) | t-value | *p* |
| Intercept | 1.84 (1.00) | 1.84 | 0.07 |
| Diagnosis | 0.27 (0.42) | 0.65 | 0.52 |
| Sex | 0.22 (0.36) | 0.61 | 0.54 |
| AgeC | -0.37 (0.02) | -19.28 | ***p*<0.001** |
| AgeC^2^ | -0.00 (0.00) | -1.74 | 0.08 |
| Diagnosis-by-ageC | -0.07 (0.03) | -2.56 | **0.01** |

Note. AgeC, mean-centered age; AgeC^2^, mean-centered age squared; Diagnosis-ageC, diagnosis-by-age-centered interaction term; LME, linear mixed-effects model; HC, healthy control; SPH, specific phobia; SE, standard error. 𝛃 is measured in years.

**Model fit statistics.**

**MAE and correlations, full sample and per diagnostic group and sex.**

Table S3. Model fit statistics, full sample, and per diagnostic group and sex.

|  | Full sample, n=1,734 | SPH, n=600 | HC, n=1,134 | Males, n=593 | Females, n=1,141 |
| --- | --- | --- | --- | --- | --- |
| MAE (SD) | 7.26 (5.41) | 7.42 (5.81) | 7.17 (5.19) | 6.90 (5.00) | 7.44 (5.60) |
| R (brain age, age) | 0.80 | 0.76 | 0.81 | 0.81 | 0.79 |
| R^2^ (brain age, age) | 0.63 | 0.58 | 0.66 | 0.65 | 0.62 |
| R (brain-PAD, age) | 0.54 | 0.52 | 0.53 | 0.50 | 0.55 |
| R^2^ (brain-PAD, age) | 0.29 | 0.27 | 0.28 | 0.25 | 0.30 |

Note: brain-PAD, brain predicted age difference; HC, healthy control; MAE, mean absolute error; R, Pearson correlation; SD, standard deviation; SPH, specific phobia.

**MAE per site and diagnostic group.**

Table S4. Mean absolute error per study site and diagnostic group.

|  | HC | | SPH | |
| --- | --- | --- | --- | --- |
| Site | **n** | **MAE (SD)** | **n** | **MAE (SD)** |
| Barcelona | 7 | 8.60 (5.80) | 22 | 7.94 (5.45) |
| Dresden CRC940C5 | 44 | 7.58 (4.97) | 52 | 10.47 (5.90) |
| Dresden SPH subtypes | 18 | 8.81 (5.36) | 39 | 7.29 (4.68) |
| FOR2107 MR | 322 | 7.19 (4.93) | 11 | 5.46 (4.78) |
| FOR2107 MS | 146 | 6.61 (5.00) | 24 | 7.36 (6.43) |
| Graz I | 26 | 8.98 (7.53) | 29 | 8.82 (5.88) |
| Graz II | 15 | 7.71 (4.67) | 16 | 7.07 (5.79) |
| Greifswald Spider Snake | 10 | 8.26 (5.28) | 10 | 10.48 (5.60) |
| Muenster Dental Phobia | 14 | 9.43 (5.84) | 12 | 6.38 (5.00) |
| Muenster SFBTRR-58 C09 | NA | NA | 58 | 7.13 (5.39) |
| Muenster Spider | 212 | 4.97 (5.22) | 22 | 6.71 (4.98) |
| Protect-AD | NA | NA | 34 | 8.73 (6.47) |
| RepSpi | 11 | 6.78 (3.56) | 8 | 3.47 (2.53) |
| SHIP | 303 | 7.22 (5.48) | 135 | 6.47 (6.40) |
| Teneriffa | 6 | 7.89 (4.77) | 26 | 6.23 (5.48) |
| Uppsala | NA | NA | 40 | 7.87 (5.03) |
| Wuerzburg SFBTRR-58 C09 | NA | NA | 62 | 7.52 (5.27) |

Note. Dresden CRC940C5: DFG Collaborative Research Centre 940, project C5; FOR2107 MR: DFG-Research Group 2107 Marburg site; FOR2107 MS: DFG-Research Group 2107 Muenster site; HC, healthy controls; MAE, mean absolute error; Muenster SFBTRR-58 C09: DFG Collaborative Research Centre Transregio 58, project C09, Muenster site; N, number; NA, not applicable; NS, not significant; Protect-AD: Providing Tools for Effective Care and Treatment of Anxiety Disorders consortium, specific phobia sample; SD, standard deviation; SHIP: Study of Health in Pomerania; SPH, specific phobia; Wuerzburg SFBTRR-58 C09: DFG Collaborative Research Centre Transregio 58, project C09, Wuerzburg site.

**Correlations per site and diagnostic group.**

Table S5. Pearson’s R and R^2^: brain age and age and brain-PAD and age, per study site and diagnostic group.

| Site | Correlations | HC | | SPH | |
| --- | --- | --- | --- | --- | --- |
|  |  | **Brain age, age** | **Brain-PAD, age** | **Brain age, age** | **Brain-PAD, age** |
| Barcelona | R | 0.78 | 0.64 | 0.40 | 0.08 |
|  | R^2^ | 0.60 | 0.41 | 0.16 | 0.01 |
| Dresden CRC940C5 | R | 0.42 | 0.37 | 0.36 | 0.44 |
|  | R^2^ | 0.18 | 0.14 | 0.13 | 0.20 |
| Dresden SPH subtypes | R | 0.49 | 0.14 | 0.60 | 0.13 |
|  | R^2^ | 0.24 | 0.12 | 0.36 | 0.02 |
| FOR2107 MR | R | 0.74 | 0.58 | 0.86 | 0.09 |
|  | R^2^ | 0.54 | 0.34 | 0.75 | 0.01 |
| FOR2107 MS | R | 0.73 | 0.57 | 0.66 | 0.70 |
|  | R^2^ | 0.54 | 0.33 | 0.44 | 0.48 |
| Graz I | R | 0.42 | 0.54 | 0.63 | 0.77 |
|  | R^2^ | 0.18 | 0.30 | 0.40 | 0.59 |
| Graz II | R | 0.59 | 0.62 | 0.68 | 0.82 |
|  | R^2^ | 0.35 | 0.39 | 0.46 | 0.67 |
| Greifswald Spider Snake | R | 0.84 | 0.69 | 0.03 | 0.22 |
|  | R^2^ | 0.70 | 0.48 | 0.00 | 0.05 |
| Muenster Dental Phobia | R | 0.55 | 0.61 | 0.71 | 0.57 |
|  | R^2^ | 0.31 | 0.37 | 0.51 | 0.32 |
| Muenster SFBTRR-58 C09 | R | NA | NA | 0.59 | 0.59 |
|  | R^2^ | NA | NA | 0.35 | 0.35 |
| Muenster Spider | R | 0.71 | 0.53 | 0.69 | 0.12 |
|  | R^2^ | 0.50 | 0.28 | 0.48 | 0.01 |
| Protect-AD | R | NA | NA | 0.70 | 0.35 |
|  | R^2^ | NA | NA | 0.49 | 0.12 |
| RepSpi | R | 0.28 | 0.10 | 0.91 | 0.67 |
|  | R^2^ | 0.08 | 0.01 | 0.82 | 0.45 |
| SHIP | R | 0.69 | 0.52 | 0.56 | 0.55 |
|  | R^2^ | 0.47 | 0.27 | 0.31 | 0.31 |
| Teneriffa | R | 0.43 | 0.81 | 0.66 | 0.64 |
|  | R^2^ | 0.18 | 0.66 | 0.44 | 0.41 |
| Uppsala | R | NA | NA | 0.58 | 0.29 |
|  | R^2^ | NA | NA | 0.33 | 0.08 |
| Wuerzburg SFBTRR-58 C09 | R | NA | NA | 0.40 | 0.52 |
|  | R^2^ | NA | NA | 0.16 | 0.27 |

Note. brain-PAD, brain predicted age difference; Dresden CRC940C5: DFG Collaborative Research Centre 940, project C5; FOR2107 MR: DFG-Research Group 2107 Marburg site; FOR2107 MS: DFG-Research Group 2107 Muenster site; HC, healthy controls; Muenster SFBTRR-58 C09: DFG Collaborative Research Centre Transregio 58, project C09, Muenster site; N, number; NA, not applicable; Protect-AD: Providing Tools for Effective Care and Treatment of Anxiety Disorders consortium, specific phobia sample; R, Pearson’s R; R^2^, Pearson’s R-squared; SHIP: Study of Health in Pomerania; SPH, specific phobia; Wuerzburg SFBTRR-58 C09: DFG Collaborative Research Centre Transregio 58, project C09, Wuerzburg site.

**Brain-PAD residuals plot.**
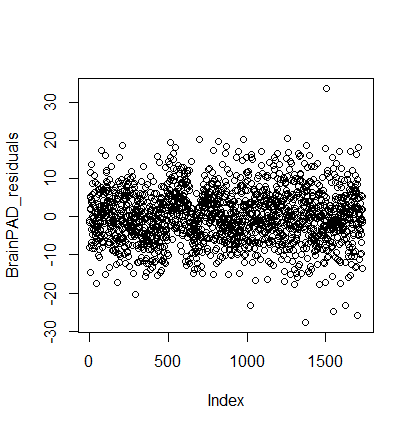

Figure S7. Brain-PAD residuals plot for primary linear mixed-effects model without interaction term. Note. BrainPAD, brain predicted age difference. Outcome variable: brain-PAD, predictor: diagnosis (specific phobia; healthy controls), random factor: study site, covariates: mean-centered age, mean-centered age^2^, sex).

**Brain age by age scatterplots.**
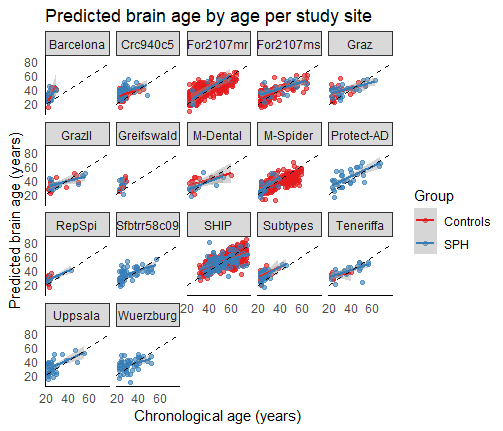

Figure S8. Brain age by age scatterplots per diagnostic group and research site. Note. Crc940c5, Dresden CRC940C5: DFG Collaborative Research Centre 940, project C5; For2107mr, FOR2107 MR: DFG-Research Group 2107 Marburg site; For2107ms, FOR2107 MS: DFG-Research Group 2107 Muenster site; M-Dental; Muenster Dental; Sfbtrr58C09, Muenster SFBTRR-58 C09: DFG Collaborative Research Centre Transregio 58, project C09, Muenster site; M-Spider, Muenster Spider; SHIP: Study of Health in Pomerania; SPH, specific phobia; Subtypes, Dresden Phobia Subtypes; Protect-AD: Providing Tools for Effective Care and Treatment of Anxiety Disorders consortium, specific phobia sample; Uppsala, Uppsala Spider Phobia; Wuerzburg, Wuerzburg SFBTRR58C09: DFG Collaborative Research Centre Transregio 58, project C09, Wuerzburg site..

**Primary linear mixed-effects model (with diagnosis-by-age interaction term) conducted in sites with HCs.**

Table S6. Between-group differences (SPH n=480 vs HC n=1,134) in brain-PAD, sites with HCs.

|  | 𝛃 (SE) | t-value | *p* |
| --- | --- | --- | --- |
| Intercept | 1.74 (1.03) | 1.69 | 0.09 |
| Diagnosis | 0.29 (0.45) | 0.66 | 0.51 |
| Sex | 0.27 (0.37) | 0.73 | 0.47 |
| AgeC | -0.37 (0.02) | -19.29 | ***p*<0.001** |
| AgeC^2^ | -0.00 (0.00) | -2.24 | **0.03** |
| Diagnosis-by-ageC | -0.05 (0.03) | -1.90 | 0.06 |

Note. AgeC, mean-centered age; AgeC^2^, mean-centered age squared; Diagnosis-ageC, diagnosis-by-age-centered interaction term; HC, healthy control; SPH, specific phobia; SE, standard error. 𝛃 is measured in years.

**Propensity score matching.**

**Matching methods and summary of balance.**

Due to the significant difference in mean age and percentage of females between SPH participants and HCs in FOR2107-MS, Muenster Spider, SHIP and Teneriffa (Table 1a), we conducted nearest neighbor propensity score matching within these sites. The summary of balance and density plots show improved balance between SPH participants and HCs after employing this procedure (Table S7, Figure S9). However, as linear age and quadratic age are controlled for in the LMEs, and both groups consisted of the same age range (22-75 years), matching was considered a supplemental analysis. In the matched dataset (SPH n=577, HC n=766), the brain-PAD effect size was slightly larger but did not significantly differ between SPH participants and HCs, while the diagnosis-by-age interaction remained significant (Table S8).

Table S7. Summary of balance for matched and unmatched data per study site.

| Study site | Variable | SPH means | HC means | Std. mean difference | Variance ratios | eCDF mean | eCDF maximum | Pair distances |
| --- | --- | --- | --- | --- | --- | --- | --- | --- |
| FOR2107 MS unmatched *(SPH n=24; HC n=146)* | **Distance** | 0.17 | 0.14 | 0.49 | 0.96 | 0.16 | 0.34 | NA |
|  | **Age** | 38.21 | 31.66 | 0.51 | 1.21 | 0.16 | 0.33 | NA |
|  | **Sex** | 0.71 | 0.66 | 0.11 | NA | 0.05 | 0.05 | NA |
| FOR2107 MS matched *(SPH n=24; HC n=48)* | **Distance** | 0.17 | 0.17 | 0.03 | 1.09 | 0.01 | 0.04 | 0.04 |
|  | **Age** | 38.21 | 37.71 | 0.04 | 1.11 | 0.02 | 0.06 | 0.13 |
|  | **Sex** | 0.71 | 0.75 | -0.09 | NA | 0.04 | 0.04 | 0.46 |
| Muenster Spider unmatched *(SPH n=22; HC n=212)* | **Distance** | 0.31 | 0.07 | 1.33 | 2.49 | 0.37 | 0.62 | NA |
|  | **Age** | 26.64 | 39.85 | -2.47 | 0.24 | 0.35 | 0.55 | NA |
|  | **Sex** | 0.86 | 0.54 | 0.95 | NA | 0.33 | 0.33 | NA |
| Muenster Spider matched *(SPH n=22; HC n=34)* | **Distance** | 0.31 | 0.29 | 0.10 | 1.18 | 0.01 | 0.18 | 0.08 |
|  | **Age** | 26.64 | 25.98 | 0.12 | 1.85 | 0.03 | 0.18 | 0.31 |
|  | **Sex** | 0.86 | 0.75 | 0.33 | NA | 0.11 | 0.11 | 0.43 |
| SHIP matched *(SPH n=135; HC n=303)* | **Distance** | 0.37 | 0.28 | 0.75 | 0.76 | 0.18 | 0.35 | NA |
|  | **Age** | 51.01 | 55.25 | -0.42 | 0.71 | 0.10 | 0.23 | NA |
|  | **Sex** | 0.79 | 0.52 | 0.65 | NA | 0.27 | 0.27 | NA |
| SHIP unmatched *(SPH n=134; HC n=213)* | **Distance** | 0.37 | 0.35 | 0.15 | 1.07 | 0.03 | 0.15 | 0.13 |
|  | **Age** | 51.16 | 52.28 | -0.11 | 0.86 | 0.04 | 0.12 | 0.48 |
|  | **Sex** | 0.78 | 0.75 | 0.07 | NA | 0.03 | 0.03 | 0.23 |
| Teneriffa matched *(SPH n=26; HC n=6)* | **Distance** | 0.87 | 0.55 | 1.82 | 0.43 | 0.37 | 0.65 | NA |
|  | **Age** | 38.00 | 25.83 | 1.11 | 2.43 | 0.34 | 0.72 | NA |
|  | **Sex** | 0.77 | 0.50 | 0.64 | NA | 0.27 | 0.27 | NA |
| Teneriffa unmatched *(SPH n=4; HC n=4)* | **Distance** | 0.73 | 0.71 | 0.07 | 0.95 | 0.02 | 0.25 | 0.14 |
|  | **Age** | 27.25 | 27.00 | 0.02 | 0.85 | 0.03 | 0.25 | 0.07 |
|  | **Sex** | 0.75 | 0.75 | 0.00 | NA | 0.00 | 0.00 | 0.00 |

Note. eCDF, empirical cumulative distribution function statistics; FOR2107 MS: DFG-Research Group 2107 Muenster site; HC, healthy control; SHIP: Study of Health in Pomerania; SPH, specific phobia; std, standardized.

**Matched age and sex density plots.**

| **FOR2107 MS** | **Muenster Spider** |
| --- | --- |
| 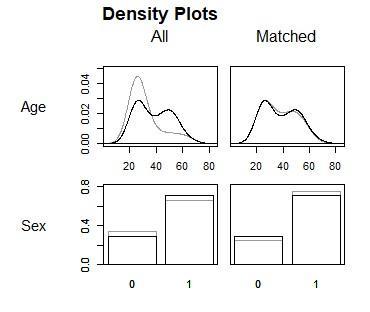 | 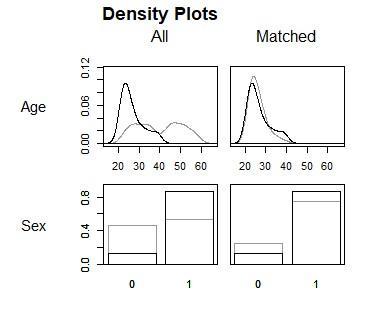 |
| **SHIP** | **Teneriffa** |
| 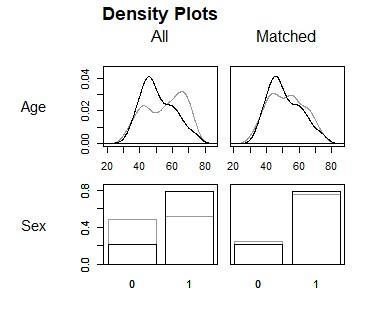 | 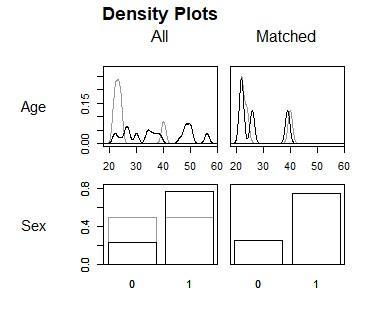 |

Figure S9. Density plots for age and sex (0 = male; 1 = female), per matched research site. Note. FOR2107 MS: DFG-Research Group 2107 Muenster site; SHIP: Study of Health in Pomerania; SPH, specific phobia; std, standardized.

**Primary linear mixed-effects model results in matched dataset.**

Table S8. Between-group differences (SPH n=577 vs HC n=766) in brain-PAD, matched dataset.

|  | 𝛃 (SE) | t-value | *p* |
| --- | --- | --- | --- |
| Intercept | 2.44 (1.03) | 2.38 | **0.02** |
| Diagnosis | 0.49 (0.46) | 1.07 | 0.28 |
| Sex | 0.44 (0.44) | 1.00 | 0.32 |
| AgeC | -0.36 (0.02) | -14.58 | ***p*<0.001** |
| AgeC^2^ | -0.00 (0.00) | -2.39 | **0.02** |
| Diagnosis-by-ageC | -0.06 (0.03) | -2.00 | **0.05** |

Note. AgeC, mean-centered age; AgeC^2^, mean-centered age squared; Diagnosis-ageC, diagnosis-by-age-centered interaction term; HC, healthy control; SPH, specific phobia; SE, standard error. 𝛃 is measured in years.

**Clinical information for study sites organized by formally diagnosed SPH and questionnaire-based SPH**

|  | SPH | Comorbidity (lifetime/current) |  | Medication use |  | AOO | STAI-T | Symptom severity centiles | BDI II |
| --- | --- | --- | --- | --- | --- | --- | --- | --- | --- |
| Site | **% Current/lifetime** | **% ANX** | **% MDD** | **% any use** | **% SSRI/SNRI** | **Mean (SD)** | **Mean (SD)** | **Mean (SD)** | **Mean (SD)** |
| Barcelona | 100.00 current | NA | NA | NA | NA | NA | 4.27 (1.55) | 6.82 (1.40) | NA |
| Dresden CRC940C5 | 100.00 current | 15.38/7.69 | 11.54 lifetime | 0.00 | 0.00 | 5.73 (3.92) | 3.17 (1.28) | 7.85 (1.32) | 1.25 (0.62) |
| Dresden SPH subtypes | 100.00 current | 6.25/18.75 | 0.00/0.00 | 6.25 | 6.25 | NA | NA | 8.38 (0.62) | 1.88 (1.78) |
| FOR2107 MR | 100.00 current | 18.18 current | 9.09/72.73 | 63.64 | 45.45 | NA | 7.27 (1.19) | NA | 4.55 (1.29) |
| FOR2107 MS | 16.67 lifetime/83.33 current | 12.50/25.00 | 4.17/58.33 | 45.83 | 12.50 | NA | 6.00 (1.53) | NA | 3.09 (1.34) |
| Graz I | 100.00 current | 0.00 | 0.00 | 0.00 | 0.00 | 13.42 (8.26) | NA | 8.69 (1.49) | NA |
| Graz II | 100.00 current | 0.00 | 0.00 | 0.00 | 0.00 | 8.06 (4.81) | NA | 8.50 (1.10) | NA |
| Muenster SFBTRR-58 C09 | 100.00 current | 0.00 | 3.57/3.57 | 43.10 | 0.00 | 6.14 (5.18) | 3.15 (1.42) | 7.76 (0.68) | 1.25 (0.62) |
| Muenster Spider | 100.00 current | 0.00 | 0.00 | 0.00 | 0.00 | NA | 4.15 (1.66) | 7.47 (1.65) | 1.23 (0.43) |
| Protect-AD | 100.00 lifetime | 73.53 lifetime | 29.41 lifetime | 0.00 | 0.00 | NA | NA | 4.26 (2.06) | 3.21 (1.67) |
| SHIP | 0.76 lifetime/99.24 current | 15.15 lifetime | 28.79 lifetime | 22.73 | 8.33 | 14.26 (11.34) | NA | NA | 2.02 (1.41) |
| Teneriffa | 100.00 current | 0.00 | 0.00 | 0.00 | 100.00 | NA | NA | 7.31 (1.12) | NA |
| Wuerzburg SFBTRR-58 C09 | 100.00 current | 0.00 | 1.61 lifetime | 40.32 | 0.00 | 7.79 (3.83) | 3.27 (1.54) | 8.00 (0.81) | 1.26 (0.51) |
| Total across all samples | 66.47/33.53 | 11.31/2.98 | 11.51/4.56 | 20.04 | 9.72 | 9.62 (8.48) | 3.83 (1.84) | 7.49 (1.70) | 1.88 (1.37) |

Table S9. Clinical information for formally diagnosed SPH participants (n=504).

Note. ANX, anxiety; AOO, age of onset; BDI, Beck Depression Inventory II; Dresden CRC940C5: DFG Collaborative Research Centre 940, project C5; FOR2107 MR: DFG-Research Group 2107 Marburg site; FOR2107 MS: DFG-Research Group 2107 Muenster site; MDD, major depressive disorder; Muenster SFBTRR-58 C09: DFG Collaborative Research Centre Transregio 58, project C09, Muenster site; NA, not applicable; Protect-AD: Providing Tools for Effective Care and Treatment of Anxiety Disorders consortium, specific phobia sample; SD, standard deviation; SNRI, serotonin and norepinephrine reuptake inhibitor; SHIP: Study of Health in Pomerania; SPH, specific phobia; SSRI, selective serotonin reuptake inhibitor; STAI-T, State-Trait Anxiety Inventory – Trait. Information was not present for all participants; Wuerzburg SFBTRR-58 C09: DFG Collaborative Research Centre Transregio 58, project C09, Wuerzburg site.

|  | SPH | Comorbidity (lifetime/current) | | Medication use | | AOO | STAI-T | Symptom severity centiles | BDI II |
| --- | --- | --- | --- | --- | --- | --- | --- | --- | --- |
| Site | **n cut-off** | **% ANX** | **% MDD** | **% any use** | **% SSRI/SNRI** | **Mean (SD)** | **Mean (SD)** | **Mean (SD)** | **Mean (SD)** |
| Dresden SPH subtypes | 23 | 4.345 current | 0.00 | 8.70 | 0.00 | NA | NA | 8.34 (0.53) | 1.68 (1.32) |
| Greifswald Spider Snake | 10 | NA | NA | NA | NA | NA | NA | NA | NA |
| Muenster Dental Phobia | 12 | 0.00 | 0.00 | 0.00 | 0.00 | NA | 3.08 (1.62) | 7.42 (1.38) | 1.36 (0.50) |
| RepSpi | 8 | NA | NA | 0.00 | 0.00 | NA | 3.00 (1.77) | 7.50 (1.31) | NA |
| SHIP | 3 | 100.00 lifetime | 0.00 | 0.00 | 0.00 | NA | NA | NA | 2.05 (1.16) |
| Uppsala | 40 | NA | NA | 0.00 | 0.00 | 5.74 (2.780) | NA | 6.78 (1.03) | NA |
| Total across all samples | 96 | 3.13/1.04 | 0.00 | 2.08 | 0.00 | 5.74 (2.80) | 2.94 (1.55) | 6.19 (2.72) | 1.45 (0.95) |

Table S10. Clinical information for questionnaire cut-off SPH participants.

Note. ANX, anxiety; AOO, age of onset; BDI, Beck Depression Inventory II; MDD, major depressive disorder; NA, not applicable; SD, standard deviation; SNRI, serotonin and norepinephrine reuptake inhibitor; SHIP: Study of Health in Pomerania; SPH, specific phobia; SSRI, selective serotonin reuptake inhibitor; STAI-T, State-Trait Anxiety Inventory – Trait.

**Primary linear mixed-effects model conducted in subgroups.**

Table S11. Between-group differences in brain-PAD (formally diagnosed SPH, lifetime and current n=384 vs HCs n=1,134) in sites with controls vs HCs.

|  | 𝛃 (SE) | t-value | *p* | Cohen’s *d* (95% CI) |
| --- | --- | --- | --- | --- |
| Intercept | 1.47 (1.08) | 1.35 | 0.18 |  |
| Diagnosis | 0.49 (0.47) | 1.04 | 0.30 | 0.06 (-0.05-0.16) |
| Sex | 0.43 (0.38) | 1.11 | 0.27 |  |
| AgeC | -0.38 (0.02) | -19.84 | ***p*<0.001** |  |
| AgeC^2^ | -0.00 (0.00) | -1.93 | 0.05 |  |
| Diagnosis-by-ageC | -0.07 (0.03) | -2.36 | **0.02** |  |

Note. AgeC, mean-centered age; AgeC^2^, mean-centered age squared; Diagnosis-ageC, diagnosis-by-age-centered interaction term; HC, healthy control; SPH, specific phobia; SE, standard error. 𝛃 is measured in years.

Table S12. Between-group differences in brain-PAD (medicated SPH n=101 vs HC n=1,134).

|  | 𝛃 (SE) | t-value | *p* | Cohen’s *d* (95% CI) |
| --- | --- | --- | --- | --- |
| Intercept | 0.49 (0.94) | 0.53 | 0.60 |  |
| Diagnosis | 1.10 (0.84) | 1.31 | 0.19 | 0.13 (0.02-0.24) |
| Sex | 0.46 (0.42) | 1.09 | 0.28 |  |
| AgeC | -0.39 (0.02) | -19.59 | ***p*<0.001** |  |
| AgeC^2^ | -0.00 (0.00) | -1.29 | 0.20 |  |
| Diagnosis-by-ageC | -0.09 (0.05) | -1.73 | 0.08 |  |

Note. AgeC, mean-centered age; AgeC^2^, mean-centered age squared; Diagnosis-ageC, diagnosis-by-age-centered interaction term; HC, healthy control; SPH, specific phobia; SE, standard error. 𝛃 is measured in years.

**Post-hoc groups split by median age.**

**Demographic and clinical information in participants split by median age.**

Table S13. Demographic and clinical information, participants ≤ 35.

|  | SPH, n=355 | HC, n=532 |
| --- | --- | --- |
| % females | 80.85^**^ | 61.09^**^ |
| Mean (SD) age | 25.87 (3.38)^**^ | 26.81 (3.87)^**^ |
| Mean (SD) Thickness | 2.59 (0.09) | 2.58 (0.09) |
| Mean (SD) AOO | 9.62(8.48) | NA |
| % Anx comorbidity (current/lifetime) | 3.38/6.20 | NA |
| % MDD comorbidity (current/lifetime) | 3.38/2.54 | NA |
| Mean (SD) STAI-T | 3.70 (1.69) | 2.70 (1.34) |
| Mean (SD) SPH symptom severity centile | 7.52 (1.45) | 1.74 (1.00) |
| Mean (SD) BDI II | 1.62 (1.15) | 1.21 (0.54) |
| % any medication use | 13.80 (n=32 NA) | 0.56 (n=24 NA) |
| % SSRI/SSNI use | 4.51 (n=138 NA) | 0.18 (n=57 NA) |

Note. ANX, anxiety; AOO, age of onset; BDI, Beck Depression Inventory II; MDD, major depressive disorder; HC, healthy control; SD, standard deviation, SPH, specific phobia; SSNI, serotonin and norepinephrine reuptake inhibitor; SSRI, selective serotonin reuptake inhibitor; STAI-T, State-Trait Anxiety Inventory – Trait. ** *p* < .001 (independent samples T-Test for mean age, and Chi-square test for % females).

Table S14. Demographic and clinical information, participants > 35.

|  | SPH, n=245 | HC, n=602 |
| --- | --- | --- |
| % females | 76.33^**^ | 56.81^**^ |
| Mean (SD) age | 48.92 (9.12)^**^ | 52.45 (10.12)^**^ |
| Mean (SD) Thickness | 2.41 (0.13) | 2.39 (0.14) |
| Mean (SD) AOO | 9.66(8.52) | NA |
| % Anx comorbidity (current/lifetime) | 1.63/15.51 | NA |
| % MDD comorbidity (current/lifetime) | 4.49/20.00 | NA |
| Mean (SD) STAI-T | 4.00 (2.21) | 2.64 (1.23) |
| Mean (SD) SPH symptom severity centile | 7.23 (2.07) | 1.59 (0.71) |
| Mean (SD) BDI II | 2.09 (1.48) | 1.28 (0.60) |
| % any medication use | 21.22 (n=4 NA) | 1.26 (n=34 NA) |
| % SSRI/SNRI use | 12.24 (n=27 NA) | 0.33 (n=3 NA) |

Note. ANX, anxiety; AOO, age of onset; BDI, Beck Depression Inventory II; MDD, major depressive disorder; HC, healthy control; SD, standard deviation, SPH, specific phobia; SSNI, serotonin and norepinephrine reuptake inhibitor; SSRI, selective serotonin reuptake inhibitor; STAI-T, State-Trait Anxiety Inventory – Trait. ** *p* < .001 (independent samples T-Test for mean age, and Chi-square test for % females).

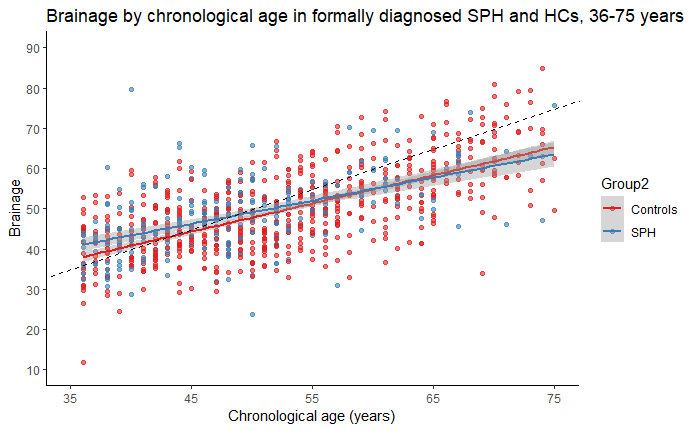

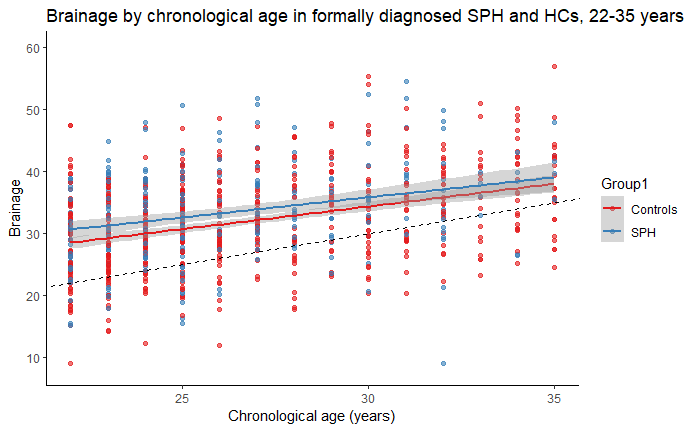
**Post hoc scatterplots of brain age by age.**

Figure S10a. Scatterplot of brain age by age in younger formally diagnosed SPH and HCs. Figure S10b. Scatterplot of brain age by age in older formally diagnosed SPH and HCs. Note. HC, healthy controls; SPH, specific phobia. Dashed line represents the line of identity (where x=y).
